# Seasonally varying effects of improved water, sanitation and handwashing interventions on Giardia infection in Bangladesh

**DOI:** 10.64898/2025.12.17.25342505

**Authors:** Pearl Anne Ante-Testard, Francois Rerolle, Mahbubur Rahman, Rashidul Haque, Shimul Das, Sarker Masud Parvez, Ayse Ercumen, Audrie Lin, Stephen P. Luby, Tarik Benmarhnia, Benjamin F. Arnold

**Author notes:** **Corresponding author:** Pearl Anne Ante-Testard; Francis I. Proctor Foundation and the Department of Ophthalmology, University of California, San Francisco, San Francisco, CA, USA;.

## Abstract

**Background:** *Giardia* is the most common enteric parasite among children in low-resource settings, causing diarrhoea and leading to prolonged infection or asymptomatic carriage. We assessed whether the effect of water, sanitation and handwashing (WSH) interventions on *Giardia* infection among rural Bangladeshi children varies with seasonal conditions.

**Methods:** We conducted a secondary analysis of the WASH Benefits Bangladesh cluster-randomized trial, with 450 clusters assigned to four arms in a 2×2 factorial design (WSH: WSH, WSH+Nutrition; no WSH: Control, Nutrition). *Giardia* infection was measured by multiplex real-time PCR in stool samples after two years of intervention. Effects were estimated by marginal treatment and assessed for heterogeneity by season when *Giardia* was measured. We also assessed heterogeneity by cumulative exposure to dry and monsoon seasons from birth to measurement age to estimate the exposure history of *Giardia*.

**Results:** *Giardia* prevalence, measured among 2773 children (median age: 30 months, range: 22-38 months), was higher in the dry seasons (32%) than in the monsoon (21%). The effect of WSH was consistent on the relative (20% reduction) and absolute scales, with slightly greater absolute reduction during the dry season (dry: −6.1%, −10.1% to −2.1%). With increasing dry-season exposure, the period of highest risk, prevalence remained consistently lower in the WSH group, with the largest differences between study groups among children with more than 17 months of dry-season exposure by age over two years.

**Conclusion:** We demonstrate how WSH provides resilience to seasonal variation in infection risk and mitigates climate-driven, seasonally varying *Giardia* transmission.

**Key Messages:**

- What your research question was We investigated whether the effect of water, sanitation and handwashing (WSH) interventions on *Giardia* infection among rural Bangladeshi children varies with seasonal conditions.
- What you found We found that *Giardia* risk varied seasonally, with improved WSH providing consistent relative and absolute reductions — slightly greater on the absolute scale during the dry season — and larger reductions among children with greater cumulative exposure to dry months, reflecting higher baseline risk and demonstrating how WSH enhances resilience to seasonally varying infection risk.
- Why it is important Since the dry and monsoon seasonal cycle is extreme in Bangladesh, this study demonstrates how WSH can mitigate seasonally varying *Giardia* infection risk.

## Background

*Giardia duodenalis* (also known as *G. lamblia* or *G. intestinalis*) is one of the most prevalent enteric parasites among children living in low-resource settings, where it contributes to both symptomatic and asymptomatic infections [1,2]. Globally, *Giardia* is a leading cause of diarrheal illness in children, following rotavirus and *Cryptosporidium* species, with over 300 million cases reported annually [3]. In areas with inadequate sanitation, giardiasis prevalence can reach 20-40%, with the highest infection rates observed in children under five years [1]. Despite its high burden, it remains one of the most neglected intestinal parasites [4]. Transmission occurs through waterborne, foodborne or person-to-person pathway. While Giardia can cause acute diarrhoea, children infected are often asymptomatic [4] with prolonged carriage that may impair growth and development in early childhood. Giardia can be prevented through water, sanitation, and handwashing (WSH) interventions, which disrupt faecal-oral transmission pathways by ensuring safe containment and disposal of faeces and by promoting hand hygiene to reduce contamination from caregivers’ hands and food [5–7].

Seasonal variation plays an important role in the transmission of enteric pathogens, including *Giardia*, but the nature of these relationships differs across settings. In urban France, rainfall and *Giardia* concentrations were weakly positively associated [8]. In rural Bangladesh, a previous analysis of the WASH Benefits Bangladesh trial reported lower *Giardia* prevalence following weeks with above-median precipitation [9], whereas a study from Mexico showed that *Giardia* incidence often peaks in the spring and summer months, particularly in warm and humid climates [10]. Seasonality may also influence interventions effects of WSH, which are a key strategy in reducing diarrhoea morbidity and mortality [11,12]. In the WASH Benefits Bangladesh trial, reductions in all-cause diarrhoea due to WSH interventions were greatest during the monsoon season [13,14]. In addition, although improved WSH reduced *Giardia* infection in the WASH Benefits trial [5], its effectiveness in mitigating climate-driven *Giardia* infection remains unclear.

Here, we assess whether the impact of WSH interventions on *Giardia* infection differs between dry and monsoon seasons, reflecting the influence of climate-sensitive exposures. We hypothesized that effect of improved WSH on *Giardia* infection could vary by season during periods of heightened environmental exposure when transmission risk is likely to be elevated.

## Methods

### Study population

We conducted a secondary analysis of the WASH Benefits Bangladesh cluster randomized trial in four rural districts in Bangladesh — Gazipur, Mymensingh, Tangail and Kishoreganj. The study design and rationale [15] and primary outcomes [11] were previously published.

The trial enrolled pregnant women who self-reported being in their first or second trimester and measured outcomes in their newborn (index) children for two years. Groups of eight mothers were organised into clusters, with each cluster spaced approximately 1 kilometre apart to minimize the risk of spill-over.

### Study design

A total of 720 clusters, grouped into blocks of eight geographically matched clusters, were randomly assigned to either a double-sized control group (C) or one of six intervention groups: improved drinking water (W), sanitation (S), handwashing (H), nutrition (N), combined water, sanitation, and handwashing (WSH), and combined WSH and nutrition (WSH+N).

This analysis focused on children from clusters that received the combined WSH interventions (WSH and WSH+N) and those from the control group (n = 360 clusters), and Nutrition arm, excluding clusters assigned to other single interventions to satisfy the consistency assumption in causal inference [16]. We analysed the interventions in a 2×2 factorial design (WSH: WSH and WSH+N, no WSH: N and control) with a focus on WSH intervention effects, as previous analyses found no evidence that the Nutrition intervention affected *Giardia* infection [5].

### Outcome

We measured *Giardia* infection status, coded as 1 for a positive test and 0 for a negative test, using multiplex real-time polymerase chain reaction (PCR), assessed ∼2.5 years after the intervention.

Trained staff distributed sterile faecal containers, instructed caregivers to collect samples from the next morning’s defecation, and retrieved them the same day. Samples were transported on ice to a satellite laboratory of the icddr,b (Dhaka), stored at −80°C, and later shipped on dry ice to icddr,b for analysis of *Giardia*. Genomic DNA was extracted from 200 mg of stool using the QIAamp Fast DNA Stool Mini Kit according to the manufacturer’s instructions and eluted in 200 µl of AE buffer [5]. Protozoa were detected by multiplex real-time PCR following a previously published protocol [17], targeting the small subunit 18s rRNA gene of *Giardia intestinalis* (62 bp, GenBank M54878). Samples were deemed positive to *Giardia* for cycle threshold (Ct) values <40 [5].

### Effect modifiers

*Season*. We defined monsoon season as the months with elevated precipitation and persistent rainfall, where the rolling 5-day average was above 10 millimetres based on previous analyses of the trial from 2012 to 2016 (May 12-October 9, 2012, April 27-October 6, 2013, May 27-September 27, 2014, April 1-September 26, 2015 and May 12-June 26, 2016) [9]. Months that fell outside monsoon seasons (coded as 0) were classified as dry season (coded as 1).

*Cumulative dry and monsoon months*. Asymptomatic carriage of *Giardia* is common, making it difficult to determine the exact timing of infection. Thus, we also used the cumulative months that a child had been exposed to the dry and monsoon period from birth up to their measurement age between 2012 and 2016 to estimate a child’s previous pathogen-specific exposure history.

### Statistical analyses

The analysis was conducted by intention-to-treat and a complete case analysis. First, we conducted descriptive statistics of the study characteristics and effect modifiers between the WSH and no WSH groups.

Second, we estimated the effect of WSH on *Giardia* by binary season (dry versus monsoon) using a generalized linear model (GLM). We used a binomial family with identity link to estimate the absolute effects (Prevalence Difference: PD) and *Giardia* prevalence, and a log link to estimate relative effects (Prevalence Ratios: PR). We accounted for clustering at block level using robust standard errors. Because the analysis followed a 2×2 factorial design, we first tested for interaction between the WSH and Nutrition interventions. As no evidence of interaction was observed (additive interaction *p*-value = 0.91, multiplicative interaction *p*-value = 0.77), we proceeded with analysing effects at the margin — the effect of each intervention (WSH) was adjusted for the other intervention and any covariates and assumed that the effect of each intervention is uninfluenced by the presence or absence of the other [18], allowing us to estimate the effects of each intervention independently [19].

We calculated the PD, PR and their standard errors [20] by estimating the marginal effect of WSH intervention on *Giardia* conditional on season and WSH (while adjusting for Nutrition). Pairwise contrasts comparing the WSH and no WSH groups were then estimated within each season stratum (dry and monsoon). We also estimated these effects using the linear combinations of the regression coefficients obtained from the regression models as a robustness check.

Third, the effect of WSH on *Giardia* prevalence was modelled as a smooth function of cumulative dry- and monsoon-month exposure using a generalized additive model (GAM) [21], conditional on cumulative season exposure and WSH and averaged over age, birth year and Nutrition. Pointwise differences between WSH and no WSH smooths were estimated [22].

### Additional and sensitivity analyses

We estimated the effect of Nutrition compared with No Nutrition to complete the 2×2 factorial analysis. We conducted sensitivity analyses that excluded the first six months of life when calculating the cumulative number of dry and monsoon months each child had experienced, excluding the earliest period of life when children were less likely to interact with their environment [23] and consume complementary foods [24].

## Results

### Study population

A total of 2273 index children were included in the analysis, of whom 985 received any form of combined WSH intervention (WSH and WSH+N) and 1288 did not (Nutrition and Control) (**Table 1**). Baseline characteristics, index child sex and potential effect modifiers were generally well balanced across study groups. Most of the children were born in 2013, accounting for nearly 90% of the participants, with fewer births in 2012 and 2014. Protozoan specimen collection was conducted in 2015 and 2016. Collection was nearly evenly distributed across 2015 and 2016, with slightly more children sampled in 2016. Overall, the children in different study groups were of similar age at the time of measurement, averaging about 2.5 years old. Most of the children were surveyed during the dry season compared to the monsoon season. The trial was conducted only in areas that were not at risk of being heavily flooded during seasonal monsoon which may partially explain more data collection in the dry season.

**Table 1.**
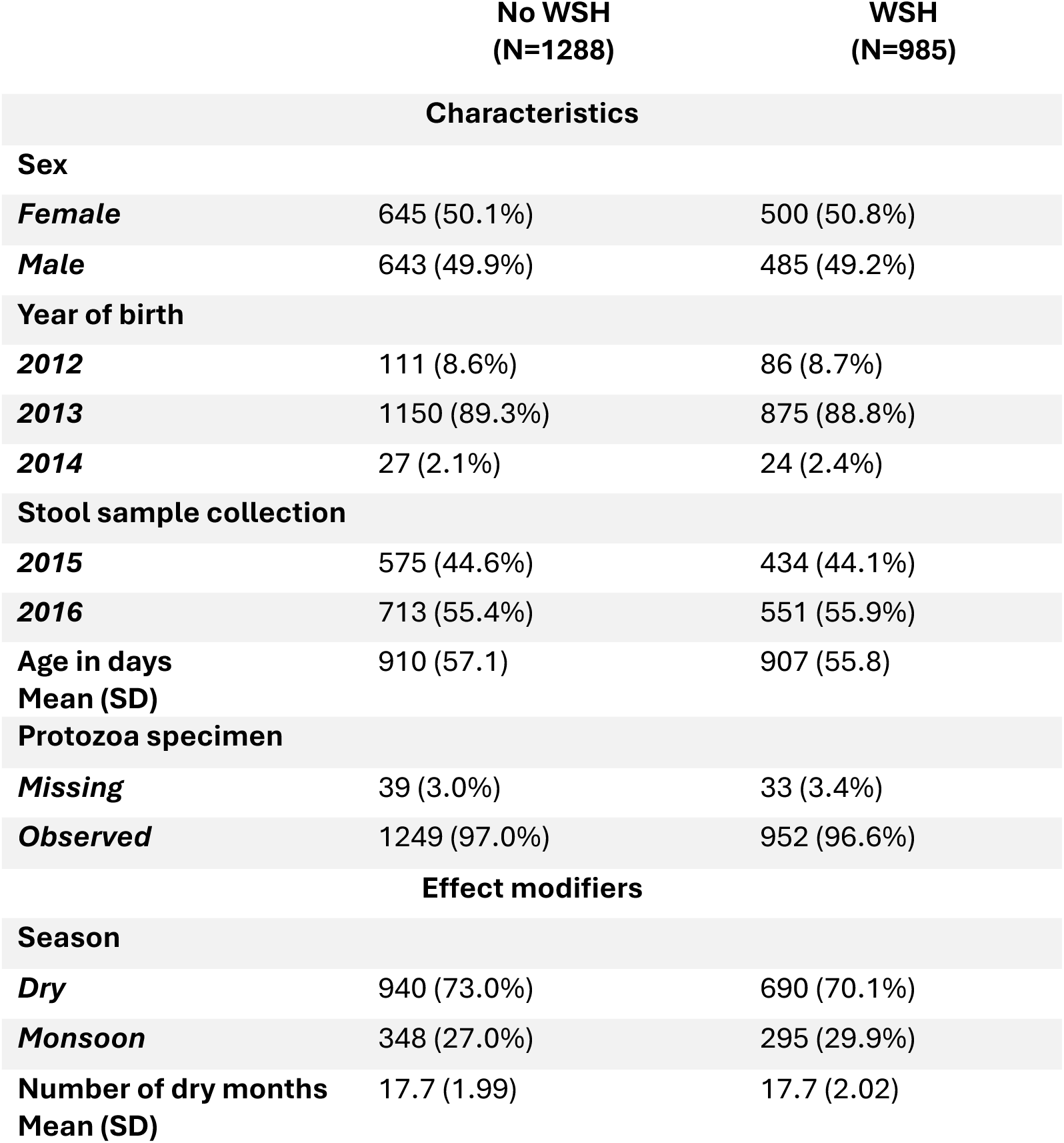
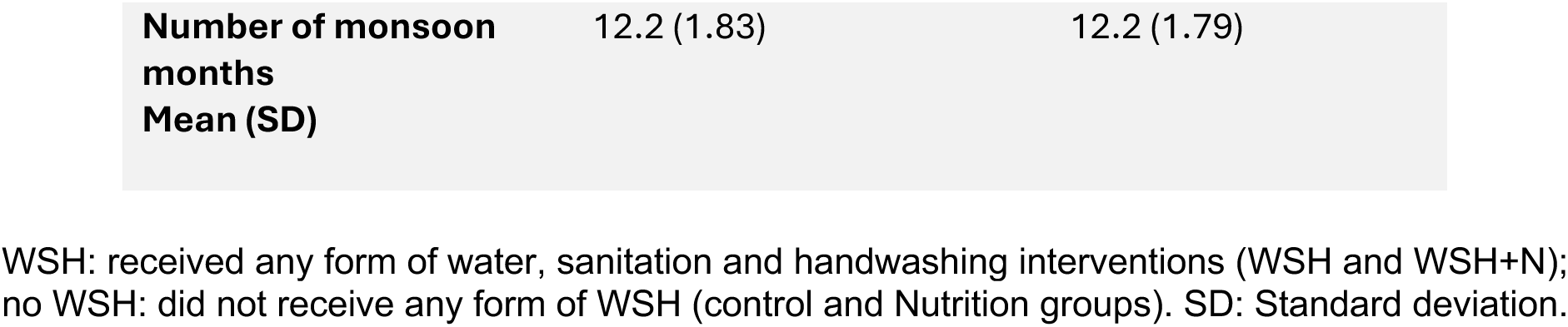
Characteristics and summary of effect modifiers between WSH and no WSH.

### Heterogeneity by exposure to the dry and monsoon seasons according to birth timing

Children were born between August 3, 2012 and May 2, 2014. Frequency of these births has occurred more during the monsoon period (**Fig. 1A**). The cumulative number of dry months experienced by children was similar between the WSH and no WSH groups (**Fig. 1B**). However, because births occurred at different times of the year, children’s cumulative exposure to dry and monsoon seasons from birth to time of measurement varied except for those born in the same day (**Fig. 1C**).

**Figure 1.**
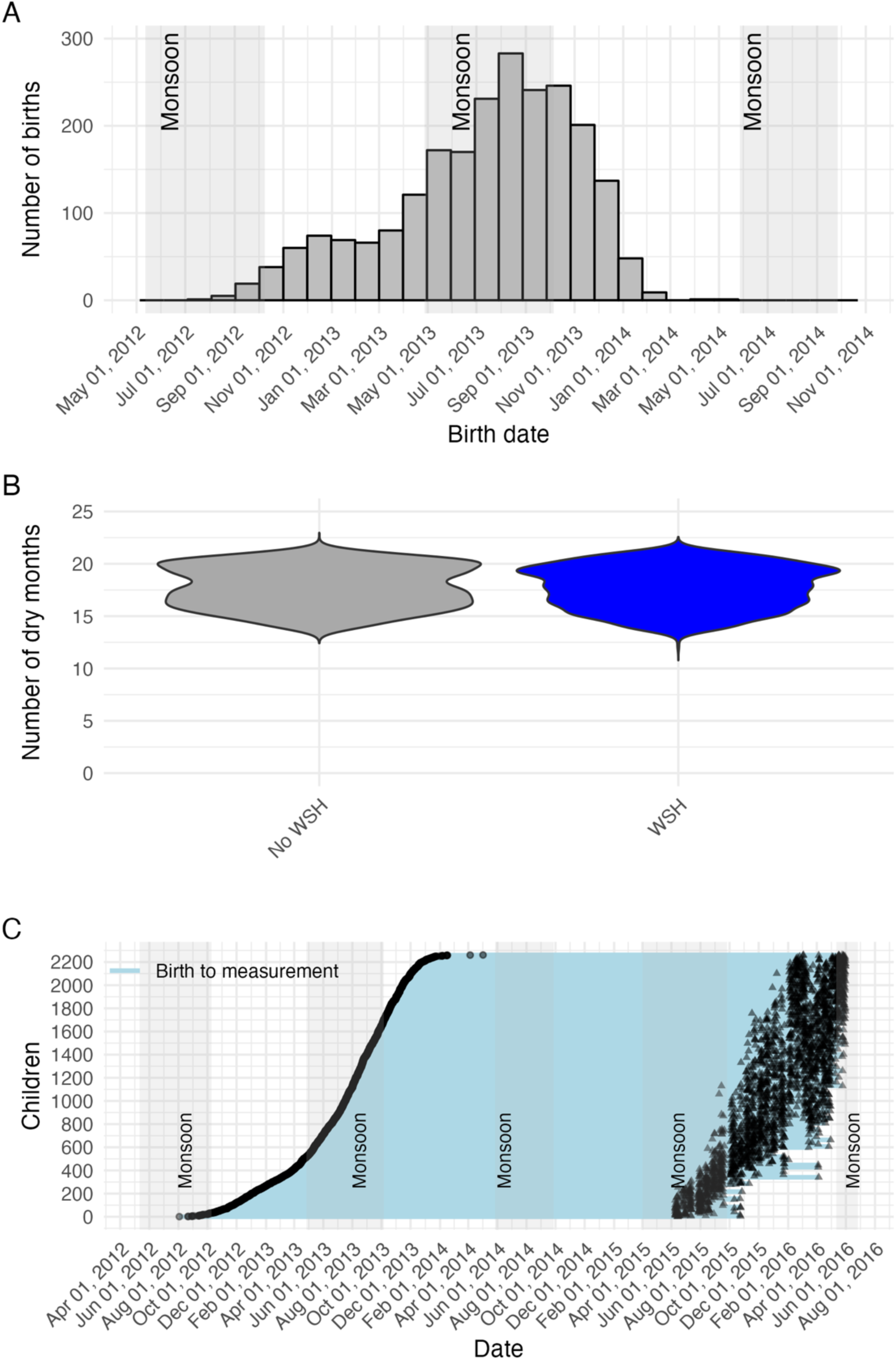
Children in the cohort experienced the dry and monsoon seasons from birth to the time of measurement. **A.** Birth dates of children included in the study with monsoon months shaded in grey. **B.** Distribution of the number of dry months by study groups **C.** Timeline from a child’s birth to measurement, with monsoon months shaded in grey to show the cumulative months of exposure to monsoon. Children are ordered by birth date.

### Effect modification by season

Overall, *Giardia* prevalence was higher in the dry season (32%) compared to the monsoon season (21%) with lower prevalence among children in the WSH group (29%, 95% confidence interval 24% to 33%) compared to no WSH (35%, 30% to 39%) (**Fig. 2A**). The prevalence during the monsoon for WSH was 19% (13% to 25%) and for no WSH was 23% (18% to 29%). The WSH interventions reduced Giardia by a similar extent both on the relative and absolute scales, with slightly greater reduction during the dry season. The absolute reduction in Giardia due to WSH appeared slightly greater in the dry season (−6.1%, −10.1% to −2.1%) than the monsoon (−4.5%, −10.6% to 1.7%) (**Fig. 2B**) with an additive statistical interaction (*p*-value for heterogeneity of 0.66). The prevalence ratio for the dry season was 0.82 (0.73 to 0.94) and for the monsoon was 0.81 (0.60 to 1.10) (**Fig. 2C**) with multiplicative statistical interaction (*p*-value for heterogeneity of 0.91).

**Figure 2.**
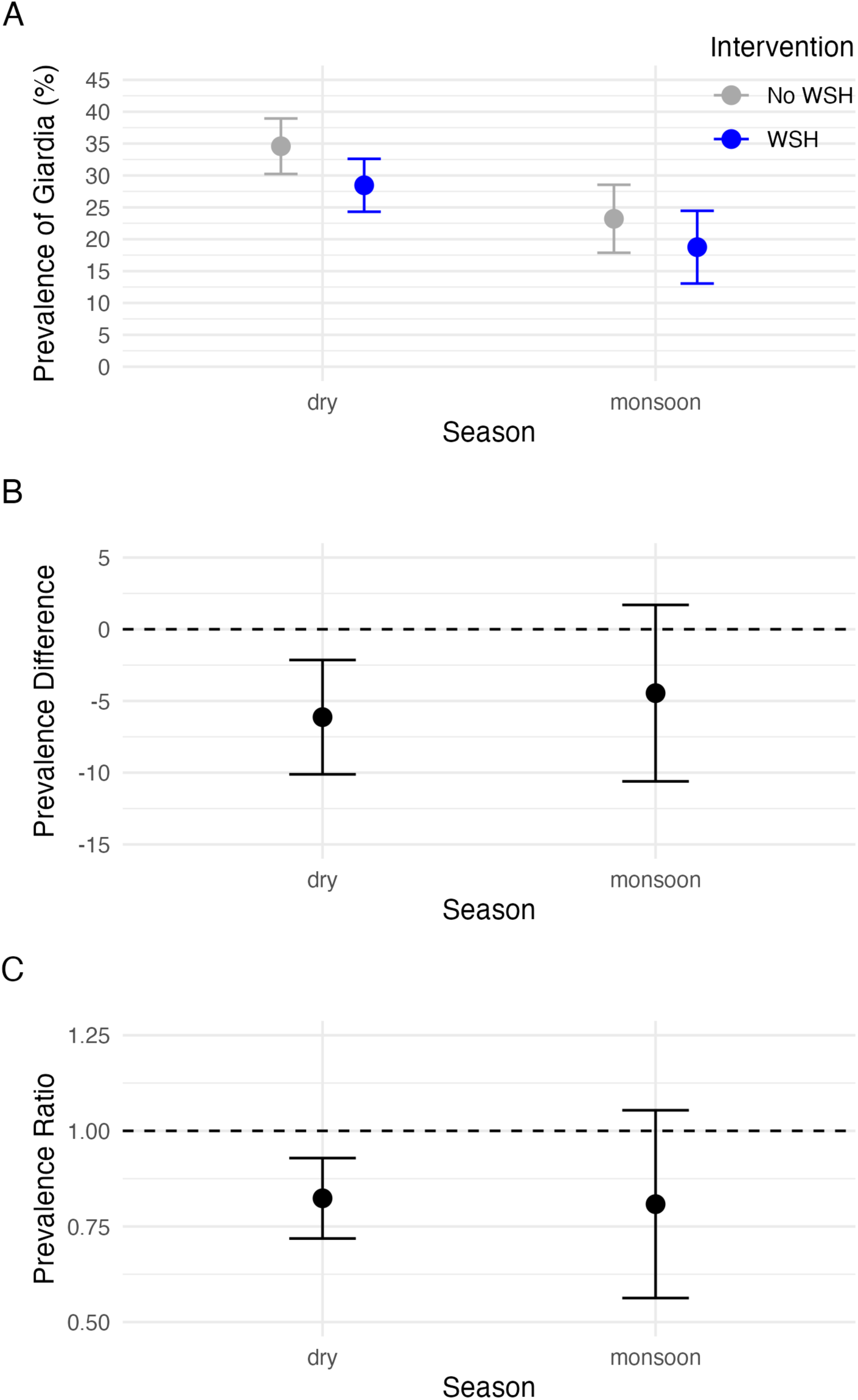
Prevalence of Giardia in the WSH and No WSH groups by season and their difference. The standard errors for the prevalence were estimated using linear combinations of the regression coefficients using the variance-covariance from the model. We accounted for clustering at cluster level using robust standard errors.

### Effect modification by cumulative exposure to dry and monsoon seasons

*Giardia* prevalence tended to increase with months exposed to the dry season for children who did not receive improved WSH. *Giardia* infection was consistently lower among children who received improved WSH compared to those who did not across the range of dry season exposure (12.2 to 21.7 months), with statistically significant differences among children with at least 17 months of dry season exposure (**Fig. 3**). When examining cumulative months of monsoon exposure, the effect of WSH was generally consistent across the range of monsoon months (**Fig. S1**). The monsoon months are not simply the inverse of dry months as children’s birth dates vary in relation to the timing of monsoon seasons.

**Figure 3.**
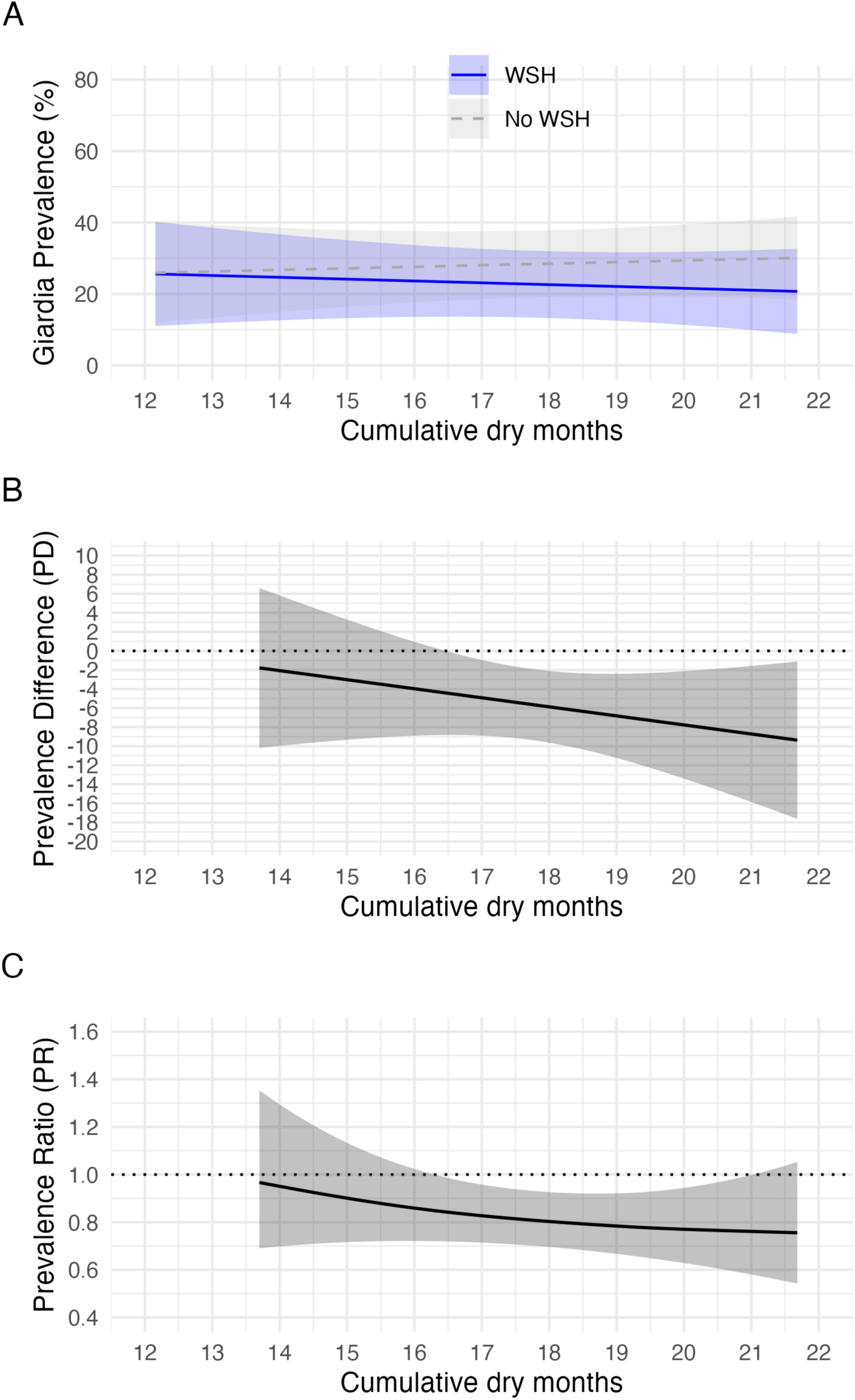
Prevalence of Giardia among children with varying exposure to dry months: A comparison between those receiving Water, Sanitation, and Handwashing (WSH) interventions and those without, within a factorial design. Adjusted for age, Nutrition and the interaction between monsoon months*WSH in the model. Used the tidymv package to estimate the smooth difference and their 95% confidence interval along the range of dry months.

### Additional and sensitivity analyses

We found that the Nutrition intervention had no effect on *Giardia* prevalence (**Fig. S2**). When repeating the analysis excluding the first six months of life, the results were consistent with the main findings, albeit the months of exposure variable is shifted due to omitting the first six months (**Fig. S3** and **Fig. S4**). *Giardia* prevalence increased with greater cumulative exposure to dry months (**Fig. S4A**), reaching up to 50% in the no WSH group by 20 months. The protective effect of WSH also increased with more exposure to dry months (**Fig. S4**).

## Discussion

We found that *Giardia* risk varied seasonally, with WSH interventions reducing *Giardia* prevalence consistently on the relative scale (20% reduction), as well as on the absolute scale with slightly greater reduction in the dry season (6% reduction). We also show that children experienced dry and monsoon seasons differently depending on their timing of birth, resulting in heterogeneous exposures to seasonal conditions from birth to the time of measurement. When accounting for cumulative exposure to dry and monsoon periods, larger reductions in *Giardia* were observed among children with greater cumulative dry-month exposure, reflecting higher absolute risk during that period. We demonstrate that cumulative exposure measures are internally consistent with seasonal stratification and highlight how improved WSH enhances resilience to seasonally varying infection risk.

We highlight the value of leveraging birth timing and cumulative climate exposures when evaluating intervention effects. By considering how children’s cumulative exposure to dry and monsoon seasons depended on their timing of birth, age, and measurement period, we could link each child’s infection outcome to their unique seasonal exposure history. This revealed that children with greater cumulative dry-month exposure slightly benefited more from improved WSH, suggesting that interventions were most protective during the dry season when infection risk was highest.

This framework estimates children’s cumulative exposure to climate conditions from birth to the time of measurement, providing a simplified representation of both the timing and duration of environmental conditions influencing infection risk. This is particularly relevant for *Giardia* which is often asymptomatic and therefore difficult to identify when infection began [4]. In this high prevalence context cumulative exposure to the dry season may also act as a proxy for prior *Giardia* exposure history. Increasing risk observed with longer dry season exposure among children without improved WSH suggests that prolonged exposure to dry conditions may facilitate sustained transmission and accumulation of infections over time.

When excluding the first six months of life of a child, a period when maternal antibodies may protect from infection [25], *Giardia* prevalence increased more with greater exposure to dry months (**Fig. S4A**). WSH intervention during the dry months of the first six months of life did not yield big effects. We also observed that the protective effect of WSH also appeared earlier, from around 13 months of cumulative dry-season exposure suggesting that children may benefit more from the intervention once maternal immunity has declined which may be due to higher *Giardia* infection and more potential to benefit from the intervention [26].

Our findings are consistent with previous studies using the WASH Benefits Bangladesh data showing that the rainy season was associated with lower *Giardia* prevalence [9]. In addition, we demonstrate consistent benefits of improved WSH and no significant benefit of the Nutrition intervention in reducing *Giardia* [5]. Findings from other settings suggest that the association between *Giardia* and environmental factors varies across contexts. A systematic review reported mixed evidence [8]. Some studies found that cyst counts increased in warm and moist conditions, while others suggested that dry or low-humidity environments may facilitate transmission [8]. A meta-analysis examining WSH interventions reported that the protective effects on diarrhoea were stronger during the dry season [27]. This aligns with our findings that WSH interventions may have the greatest impact during the dry period. However, our findings contradict with a previous analysis using the same data showing greater diarrhoea reductions during wet seasons [13,14] highlighting that the direction of seasonal effects may depend on the etiological agent (e.g., all cause diarrhoea versus bacteria versus protozoa versus virus).

Both heavy rainfall and prolonged dry periods can influence enteric pathogen transmission, albeit through distinct pathways. Intense precipitation or monsoon rains may overwhelm sanitation and drainage infrastructure, mobilize animal and agricultural waste into surface water used for recreation or irrigation, and contaminate groundwater sources [28,29]. Rainfall also facilitates the transport of pathogens across environmental compartments. Conversely, extended dry conditions and extreme temperature can elevate infection risk by concentrating pathogens in diminishing or stagnant water sources, reducing water availability for hygiene, and increasing reliance on contaminated supplies [30]. Drought conditions can lead to the accumulation of enteric pathogens in surface waters, which may subsequently be flushed and diluted during rainfall events [8,31]. Consistent with this mechanism, *Cryptosporidium* oocyst concentrations have been shown to correlate negatively with precipitation, not because rainfall introduced additional oocysts, but because it diluted those already present [8,32]. A similar process may underlie *Giardia* transmission, whereby pathogen concentrations and exposure risk rise during dry periods and decline with increased rainfall and water turnover during the monsoon. In our study, this mechanism may be reflected in the observed decrease in *Giardia* prevalence with greater cumulative exposure to monsoon months (**Fig. S2A**) and the corresponding increase with longer exposure to dry months (**Fig. 3A**) in the no WSH group. These findings suggest that improved WSH conditions may provide resilience during dry periods, when concentrated contamination and reduced water availability elevate transmission risk.

Our study had several limitations. First, the trial was conducted in areas that were not highly prone to flooding during the seasonal monsoon which may have led to underestimation or overestimation of the effects of WSH on *Giardia* compared with outcomes that might be observed in more flood-prone settings. Second, the study only measured children ages 22-38 months at the time of measurement and so did not characterize risk at other ages. Third, we defined season using only the monsoon periods of 2015-2016, which may not fully reflect broader climate variability or interannual fluctuations. To address this, we also created a continuous measure of cumulative exposure to dry and monsoon months from birth to measurement, which may better capture these dynamics. Future studies examining different enteric pathogens and a wider range of climate exposures could provide deeper insight into effect heterogeneity across pathogens and seasonally varying conditions.

The study also had several strengths. We analysed the effect of WSH on *Giardia* within a 2×2 factorial design which enhanced statistical power [18]. In addition, we incorporated cumulative exposure to dry and monsoon seasons providing a proxy for *Giardia* exposure history while also accounting for climate-related heterogeneity.

In conclusion, we demonstrate that *Giardia* risk varies seasonally, with improved WSH providing consistent relative risk reduction in both seasons and the largest absolute reductions during the dry season, the period of elevated infection risk. This finding underscores the importance of WSH interventions for strengthening population resilience to seasonally and climate-driven fluctuations in disease transmission, particularly in climate-sensitive and low-resource settings.

## Ethics approval

The protocol of the original study (Clinical Trial Registration NCT01590095) was approved by the Ethical Review Committee at the International Centre for Diarrhoeal Disease Research, Bangladesh (PR11063), the Committee for the Protection of Human Subjects at the University of California, Berkeley (2011-09-3652), the Institutional Review Board at Stanford University (25863) and at the University of California, San Francisco (22-36722). Primary caregivers provided written informed consent.

## Acknowledgements

We are grateful to Mohammad Zahidur Rahman and Mohammad Alimojjaman for supervising the data and specimen collection.

## Author contributions

P.A.A.-T. and B.F.A. conceptualised the study. B.F.A. and P.A.A.-T. acquired the study funding. P.A.A.-T. conducted the analysis with input and guidance from B.F.A. P.A.A.-T. constructed the table and figures with input from B.F.A. P.A.A.-T. wrote the initial draft of the manuscript with input and conceptual guidance from B.F.A. All authors contributed to the subsequent revisions. All authors read and approved the manuscript.

## Conflict of interest

The authors declare no competing interests.

## Funding

The WASH Benefits Bangladesh trial was funded by the Bill & Melinda Gates Foundation (OPPGD759) with additional support for this analysis from the National Institute of Allergy and Infectious Diseases (R01AI166671 and R03AI188012 to B.F.A.).

## Data availability

The de-identified data and codes are available through the Open Science Framework (OSF, https://osf.io/bc4xk/) [33].

## Notes

### Competing Interest Statement

The authors have declared no competing interest.

### Clinical Trial

Clinical Trial Registration NCT01590095

