## Supplementary Material for "Seasonally varying effects of improved water, sanitation and handwashing interventions on Giardia infection in Bangladesh"

**Figure S1**. **Prevalence of Giardia among children with varying exposure to monsoon months: A comparison between those receiving water, sanitation, and handwashing (WSH) interventions and those without, within a factorial design.** Adjusted for age, birth year, Nutrition and the interaction between monsoon months and WSH in the model.

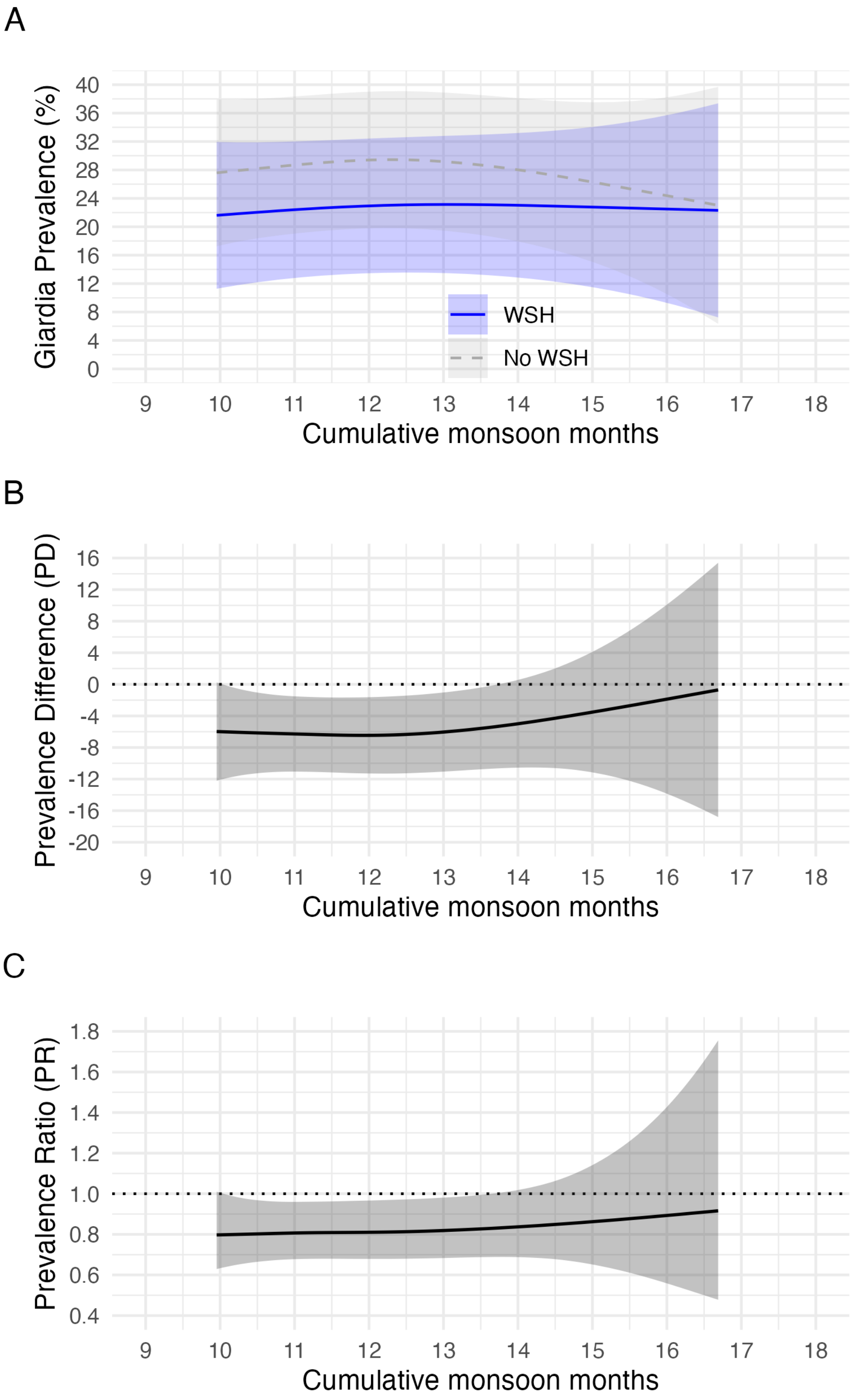

**Figure S2. Prevalence of Giardia among children with varying exposure to dry months: A comparison between those receiving nutrition interventions and those without, within a factorial design.** Adjusted for age, birth year, water, sanitation and handwashing (WSH) group and the interaction between dry months and Nutrition in the model.

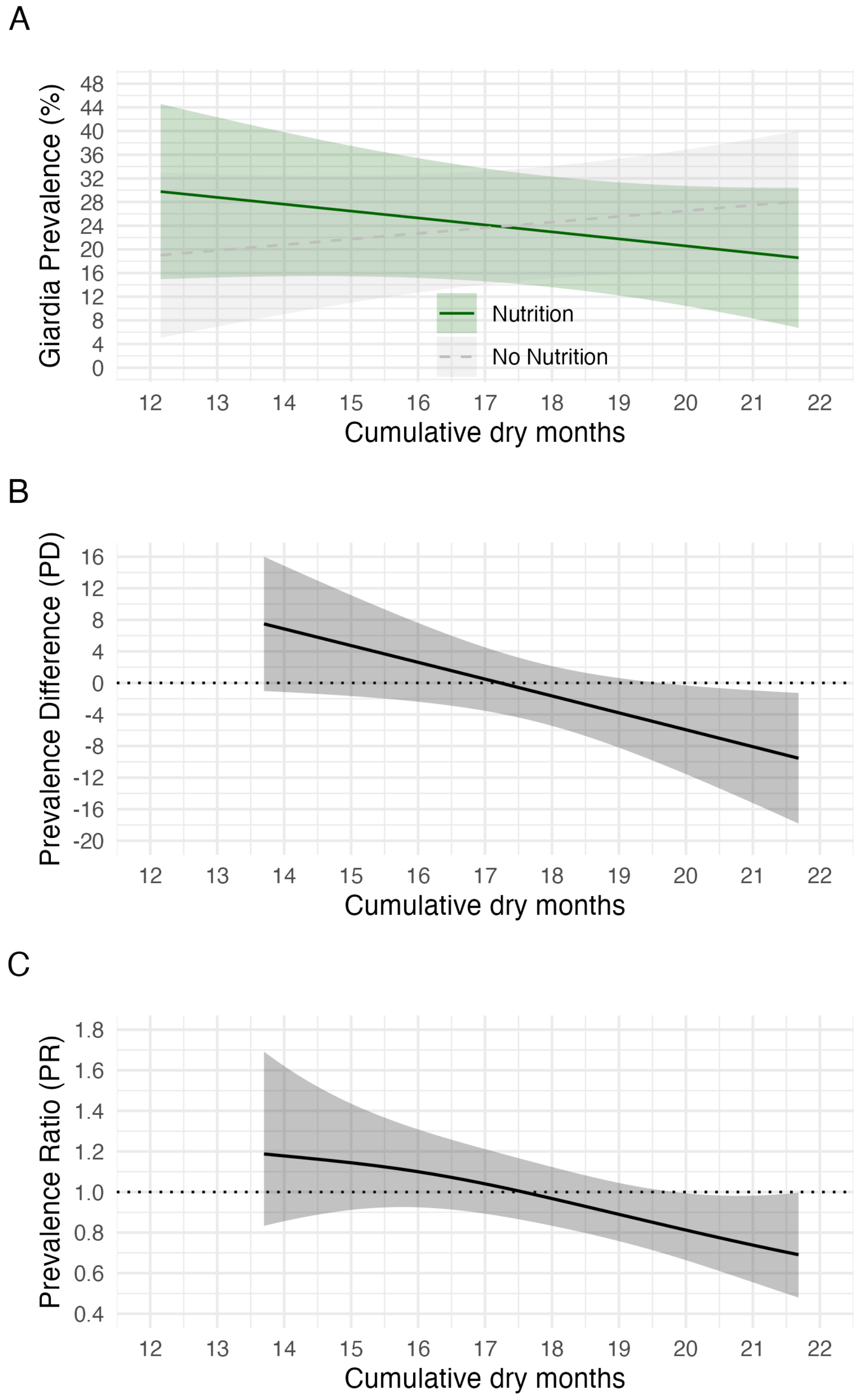

**Figure S3. Children in the cohort experienced the dry and monsoon seasons from six months after birth to the time of measurement. A.** Six months after birth of children included in the study with monsoon months shaded in grey. **B.** Distribution of the number of dry months by study arms. **C.** Timeline from six months after birth to measurement, with monsoon months shaded in grey to show the cumulative months of exposure to monsoon. Children were ordered by birth date.

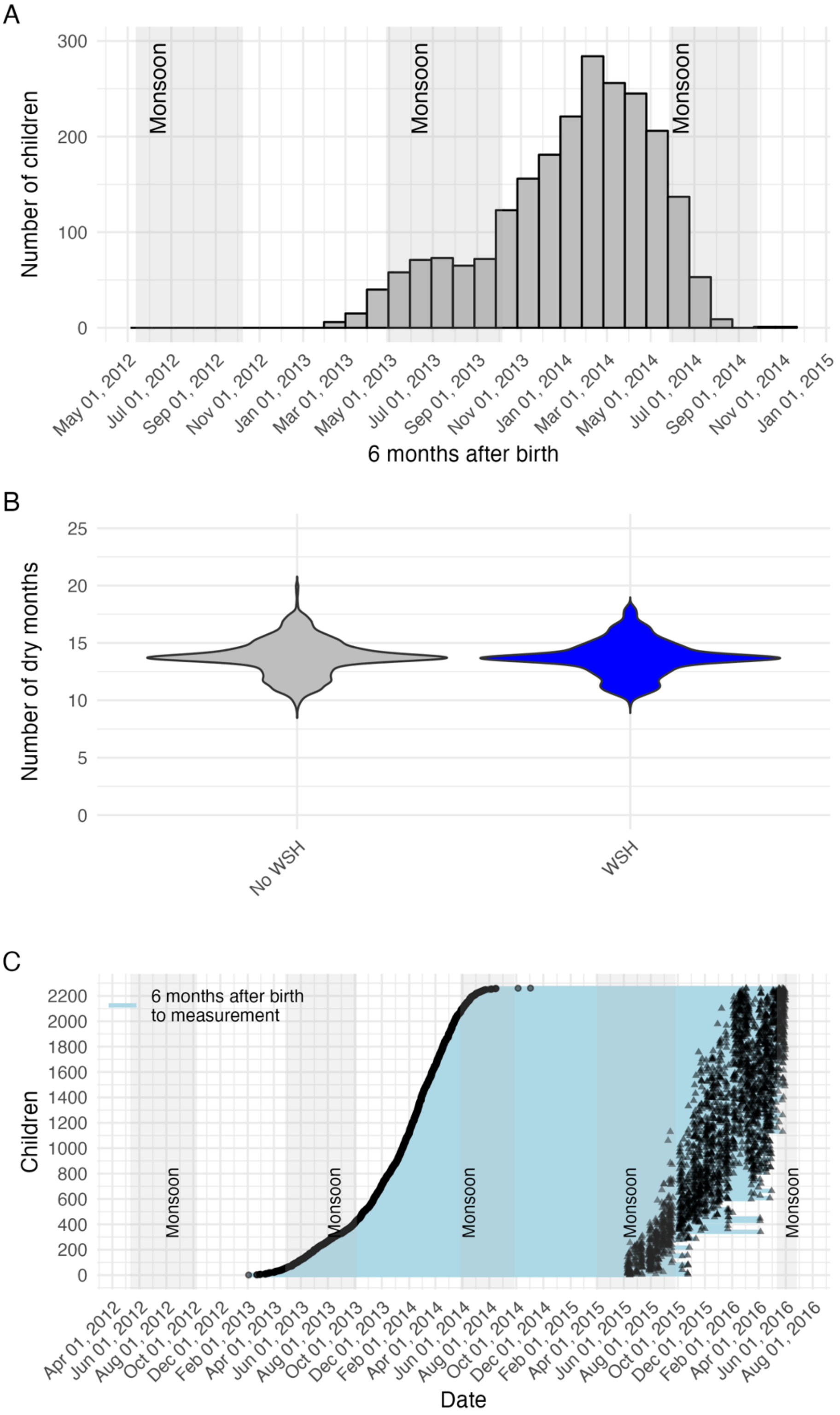

**Figure S4**. **Sensitivity analysis excluding the first 6 months of life to estimate the number of monsoon months a child had experienced.** Prevalence of Giardia among children with varying exposure to dry months: A comparison between those receiving water, sanitation, and handwashing (WSH) and Nutrition interventions and those without, within a factorial design. Adjusted for age, birth year, nutrition intervention and the interaction between monsoon months and WSH in the model.

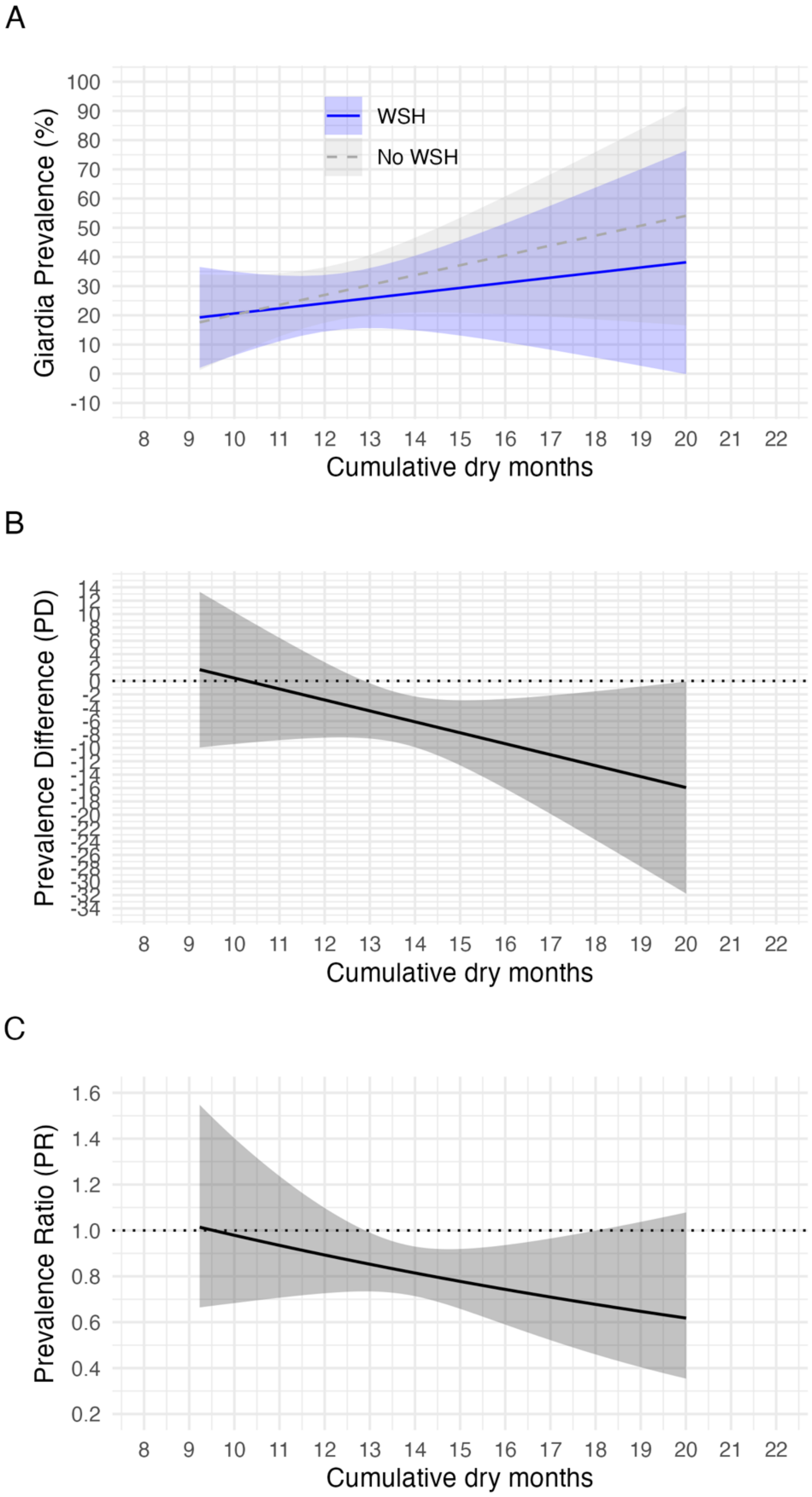
